# Infusing behavior science into large language models for activity coaching

**DOI:** 10.1101/2023.03.31.23287995

**Authors:** Madhurima Vardhan, Narayan Hegde, Deepak Nathani, Emily Rosenzweig, Alan Karthikesalingam, Martin Seneviratne

**Affiliations:** Google Research, Bangalore, India; Verily Life Sciences, San Francisco, USA; Google Health, London, UK

## Abstract

Large language models (LLMs) have shown promise for task-oriented dialogue across a range of domains. The use of LLMs in health and fitness coaching is under-explored. Behavior science frameworks such as COM-B, which conceptualizes behavior change in terms of capability (C), Opportunity (O) and Motivation (M), can be used to architect coaching interventions in a way that promotes sustained change. Here we aim to incorporate behavior science principles into an LLM using two knowledge infusion techniques: coach message priming (where exemplar coach responses are provided as context to the LLM), and dialogue re-ranking (where the COM-B category of the LLM output is matched to the inferred user need). Simulated conversations were conducted between the primed or unprimed LLM and a member of the research team, and then evaluated by 8 human raters. Ratings for the primed conversations were significantly higher in terms of empathy and actionability. The same raters also compared a single response generated by the unprimed, primed and re-ranked models, finding a significant uplift in actionability from the re-ranking technique. This is a proof of concept of how behavior science frameworks can be infused into automated conversational agents for a more principled coaching experience.

**Institutional Review Board (IRB):** The study does not involve human subjects beyond the volunteer annotators. IRB approval was not sought for this research.

## 1. Introduction

It is estimated that 81% of adolescents and 27% of adults do not achieve the levels of physical activity recommended by the World Health Organization (WHO) (1). A sedentary lifestyle is associated with long term adverse health outcomes, ranging from cardiovascular disease and diabetes to mental health problems and cognitive decline (2). A 2022 report found that progress toward these goals has been slower than expected and highlighted digital health tools as a particular opportunity area (3).

Numerous smartphone nudging tools have been designed to promote physical activity (4; 5). These interventions are low-cost and highly scalable relative to human fitness coaches, with promising early evidence (6; 7; 8; 9). One randomized controlled trial of a digital walking coach found short-term improvements in physical activity (10). However, in an era of notification overload, there is also a risk of desensitization and alert fatigue if the nudge strategy is not well designed.

Automated conversational agents offer an opportunity to create interactive dialogue, with widespread applications in e-commerce, home automation and healthcare (11; 12). Health and Fitness coaching is emerging as a promising use case for these conversational agents (13; 14; 15; 16). However, most traditional systems are limited in their degree of personalization and persuasiveness because they depend on rule-based nudge engines with static message content rather than adaptive conversational agents that can mimic realistic dialogue from a human coach (17).

Large language models (LLMs), such as GPT-3 (18), PaLM (19), Gopher (20) and LaMDA (21), excel in natural language generation with greater expressivity and versatility compared to rule-based chatbots. To date, use of LLMs in the health and fitness space has been limited, however interest is growing rapidly following the release of LLMs tailored to biomedical tasks (22). A major challenge in using LLMs in health care is how to ensure the model is personalized and adaptive while still remaining consistent with evidence-based practice and within safety guardrails (23). Activity coaching relies on complex interpersonal dynamics where the coach builds rapport with the trainee, provides motivation, helps to overcome pre-existing patterns of behavior, etc.-which are not explicitly optimized in LLMs (24). Knowledge infusion refers to the integration of established knowledge or practice into a model. In principle this is often achieved via finetuning on a task-specific dataset (25). The disadvantage of finetuning in the coaching domain is that it requires coaching transcripts, which are difficult to obtain. Finetuning has also been shown to diminish the few-shot performance of a pretrained LLM with in-context prompts - i.e. over-specialization of the model (26). Knowledge infusion is an active area of research and many other methods exist including customizing training objectives (27), reinforcement learning with human feedback (28; 29), in-context learning via prompt engineering or priming (30; 31) and many associated prompt design variants (32; 33; 34; 35). There have also been numerous strategies to ensemble knowledge infusion techniques, including post-hoc re-ranking or summarization of model outputs to further align the model with the task of interest (36; 37). Customizing knowledge infusion strategies for the health care domain remains an area of active research. Here we propose two simple in-context learning methods to infuse behavior science principles into LLMs without the requirement for finetuning or reinforcement learning.

Coaching in the context of physical activity ranges from delivering tailored products that serve elite athletes, to creating motivational tools that support inactive users to become fitter through a progressive and personalised programs. Our LLM is designed to target latter use case to help users lead more active lifestyle using behavioral nudges and resolving barriers through conversations.

Behavioral science offers theoretical frameworks to help understand the factors influencing human behavior and design effective behavior change interventions for a given context. COM-B is a well-known framework which conceptualizes behavior change along three axes: Capability (the psychological and physical skills to act); Opportunity (the physical and social conditions to act); and Motivation (the reflective and automatic mental processes that drive action) (38). Behavioral science can be useful to guide the design of automated nudging systems for habit formation (39).

We extend the PACE (16) work on designing automated physical activity coaching engine based on an analogous behavior science framework called Fogg’s Behavior Model. A rule-based automated nudging agent based on this model had comparable outcomes to human coaches in terms of user step count and engagement. In this study, we extend findings of the PACE study by connecting the strengths of a behavioral science rule-based model with the conversational versatility of an LLM. The goal is to address the broader question of how behavior science principles might guide or constrain conversational LLMs. Specifically, we make use of priming and dialogue re-ranking. These are both lightweight techniques that do not require additional model retraining or finetuning. Overall, the key contributions contributions of this study are as follows:

1. Defining evaluation metrics for LLM conversations in the activity coaching domain
2. Introducing two different approaches to behavioral science knowledge infusion: coach phrase priming and dialogue re-ranking
3. Evaluating the benefit of knowledge infusion relative to an unprimed LLM using quantitative and qualitative approaches

## 2. Methods

The following sections outline the datasets, language modeling techniques and evaluation methods used.

### 2.1. Data

The previous PACE study dataset was re-purposed for this analysis (16). Specifically, this dataset was used to construct the example coaching phrases used in the behavior science priming, create training data for finetuning BERT user and coach statement classifiers and to select the user queries (initial user responses) in simulated conversations for evaluation. This dataset consists of dialogue transcripts between fitness coaches and subjects, generated from real coaching interactions across various activity habit formation related issues. In this Wizard-of-Oz study design, consented subjects were randomized to coaches or coaches using a FBM assistant that suggested example responses based on behavior science using a rule-based engine. The dataset included 520+ conversations from 33 participants over 21 days. A total of 6 independent annotators labeled these conversations as one of Motivation, Capability and Opportunity. Both user and coach statements where separately annotated with presence or absence of each of these three themes. Data collection and annotation protocol is described in detail in (16).

### 2.2. Language models

The Language Models for Dialog Applications (LaMDA) pretrained LLM was used as the primary architecture (21), with no further finetuning. LaMDA is a decoder-only transformer architecture with 64 layers, used here in its 137 billion parameter configuration. We used the following LaMDA hyperparameters: temperature 0.9; maximum token length 1024, top k (controls sampling diversity) 40. LaMDA has an option to provide context alongside the LLM prompt - this was how the coach phrase priming was conducted. LaMDA also provides top-k outputs, which were used in the re-ranking (see below).

### 2.3. Coach phrase priming

Coach phrase priming was performed by inputting 30 example coach nudges as context to the LLM prior to the prompt. The 30 nudges were selected from the data in the PACE study - specifically the 10 most common coach responses in each of the three behavior science categories of interest: C/O/M. Details regarding coach phrase selection and priming method are described in section 1 of supplementary paper. For example, the Capability category included activity planning and barrier conversations; and Opportunity included social engagement conversations and activity planning; and Motivation included congratulations and positive affirmation; [ref]. The order of the 30 nudges was randomized. The priming prompts are shown in Table 1.

**Table 1:**
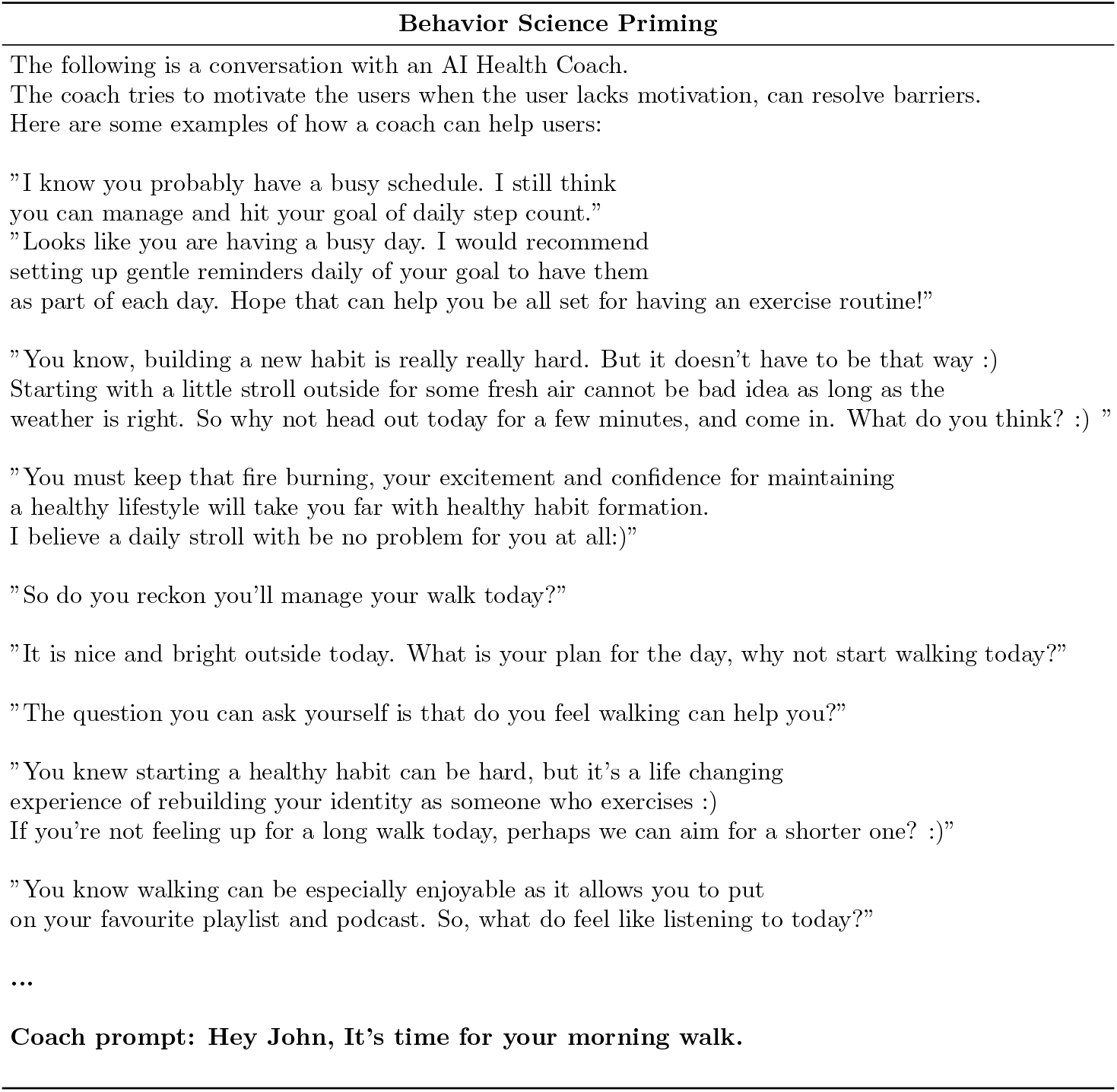
LLM prompts used in coach phrase priming.

### 2.4. Simulated dialogue

The following LLM configurations were compared via simulated conversations with a single member of the research team:

1. Unprimed (trigger prompt only)
2. Coach-primed (30 example nudges provided as LLM context)

All conversations begin with the trigger prompt: *Hey John, It’s time for your morning walk*. The subsequent user responses were sampled from a set of 9 user statements, with 3 each designed to evoke a low Motivation, low Capability and low Opportunity (user queries are included in the Supplementary Materials table 2). An example user statement with low opportunity was: *I am super busy with work today. I have chores to do in the morning and work meetings after that*..

**Table 2:**
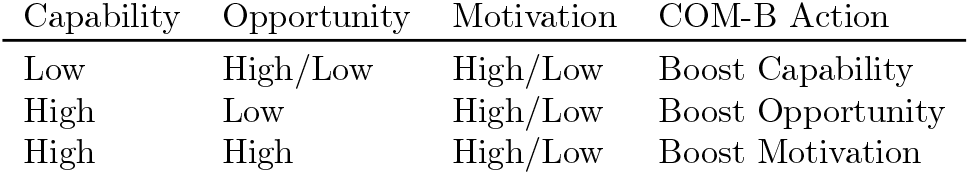
COM-B policy to select nudge theme based on C/O/M values derived from the user statement classifier.

This culminated in a total of 18 transcripts: 9 each for the unprimed and primed LLMs. The conversations were continued with dialogue between the LLM and the human interlocutor (researcher). The conversations were terminated at a natural breakpoint at the discretion of the researcher. Any follow up questions to the LLM response were added appropriately to continue the conversation on the original topic until a logical end was reached. Additional example transcripts are contained in the Supplementary Materials.

### 2.5. Constraining LLM responses using a COM-B classifier

In order to further constrain or guide the LLM to provide nudges based on COM-B principles, we trained two classifiers to assess C/O/M levels:

1. User statement classifier: Given a user statement sentence, the user-query classifier assigns a high vs low value for each of the capability, opportunity and motivation(COM) dimensions (multi-label classification).
2. Coach statement classifier: Given a shortlist of 15 top LLM outputs, the coach response classifier maps each response to either C, O or M (multi-class classification).

The classifiers were designed as follows. The input string (could be multiple sentences) was embedded using a BERT-base model with the final layer finetuned over either a multi-label head (user statement classifier) or 3 separate C/O/M heads (coach statement classifier). Models were optimised with a crossentropy loss. Separate user and coach classifiers were trained using samples of 432 user statements and 531 coach statements from the PACE study, manually annotated with C/O/M status. These datasets were split 70:10:20 across train, validation and test splits. Weights were not shared between the user and coach models.

**Figure 1:**
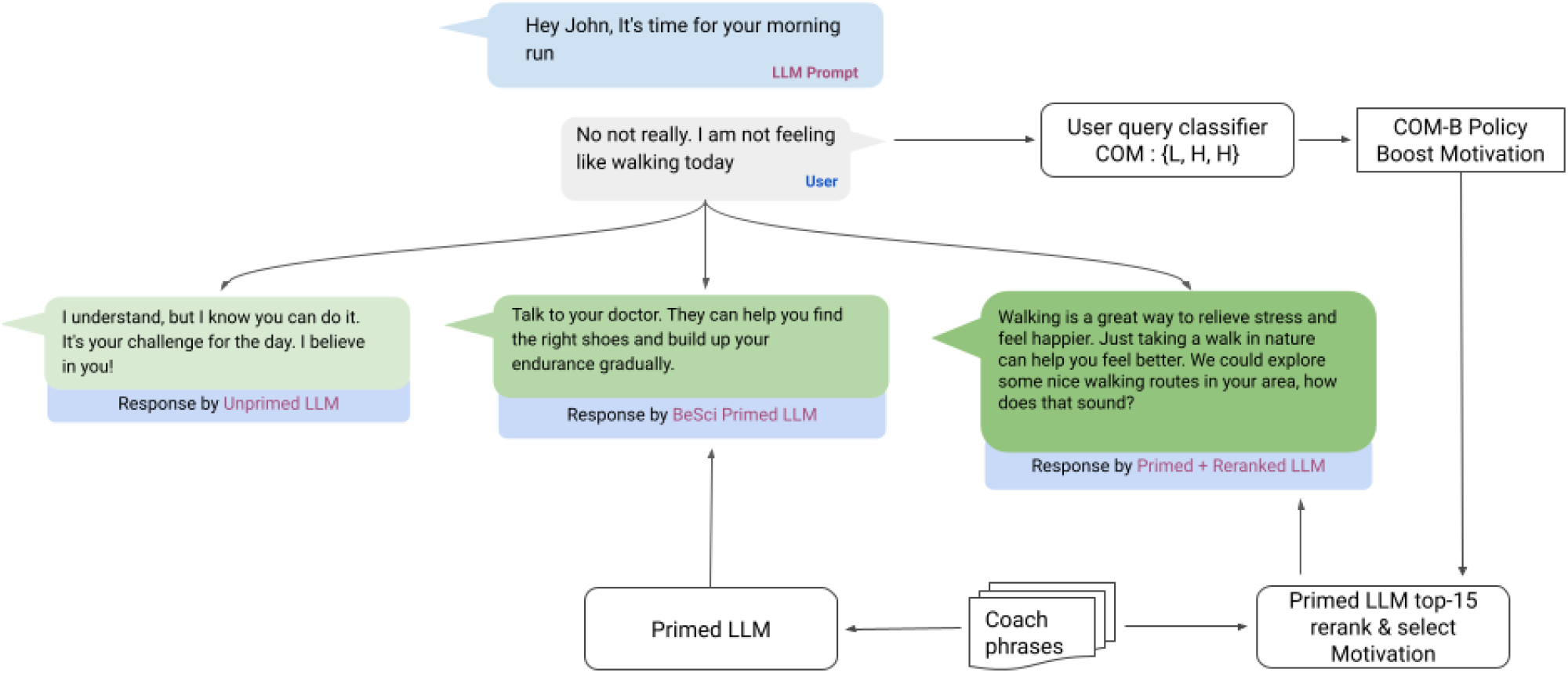
Comparison of example conversations with unprimed, coach-primed and primed+reranked LLMs.

### 2.6. Simulated dialogue with re-ranking

The simulated conversation experiment was repeated with the primed LaMDA model, using the above classifiers to align the coach response to the inferred user need. For the 9 coach-primed LaMDA transcripts above, a single user statement was manually selected as the most representative of the user’s behavioral need.

The selected text was input into the user statement classifier to identify the C/O/M need. The same user text was input into the coach-primed LaMDA model to generate the top 15 candidate responses. These 15 responses were then separately run through the coach statement classifier to generate a likelihood score across each C/O/M category. The coach action was aligned based on the user’s inferred C/O/M need based on the rules in Table 2 (i.e. the statement with the highest score in the desired coach action was chosen).

In addition, we conducted an ‘oracle’ experiment where the user response was manually categorized into C/O/M need and the corresponding coach-primed output was chosen.

Two manual review exercises were then conducted:

1. Comparing the coach-primed output to the classifier re-ranked output; and
2. Comparing coach-primed with the oracle re-ranked output. Note that in both these review exercise, only a single coach response was being adjudicated rather than an entire conversation as previous.

### 2.7. Evaluation attributes

An evaluation framework was defined based on four key attributes of an LLM-based fitness coach: actionability, realism, motivation and empathy. Coupled with a global assessment of coaching quality, these attributes informed the design of the quantitative and qualitative review detailed below. Table 3) shows how these attributes align with published evaluation frameworks for coaches (40) and for LLMs (21; 41).

**Table 3:**
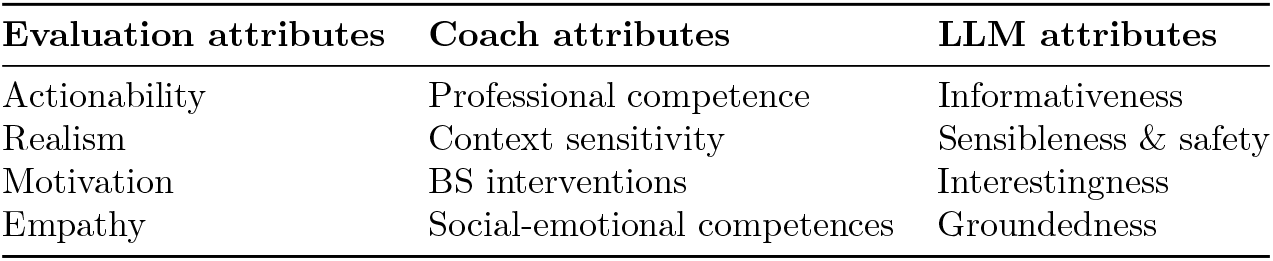
Evaluation attributes cross-referenced with established attributes of coaches and LLMs.

| Evaluation attributes | Coach attributes | LLM attributes |
| --- | --- | --- |
| Actionability | Professional competence | Informativeness |
| Realism | Context sensitivity | Sensibleness & safety |
| Motivation | BS interventions | Interestingness |
| Empathy | Social-emotional competences | Groundedness |

### 2.8. Quantitative review

For each architecture, the unprimed versus coach-primed transcripts generated from the same starting prompt were compared in a pairwise manner. The conversations were evaluated based on the following quantitative attributes: average length of reply, number of conversational turns, user sentiment at conversation end, presence of questions in the coach dialogue and use of coaching-specific words (‘goal’, ‘health’, ‘routine’, ‘recover, ‘challenge’, ‘workout’, ‘training’, ‘rest’). The results for unprimed versus primed LLMs were compared using a two sided t-test.

### 2.9. Qualitative review

8 independent reviewers were selected to adjudicate the transcripts. Reviewers were blinded to the manner of LLM priming (naive vs BS) and Re-Ranked LLM variations (naive vs primed vs re-ranked LLM). Raters completed a structured survey with Likert scale responses for the same pairwise comparisons of naive-primed and coach-primed transcripts as above. Questions evaluated the following attributes of the conversation: actionability, realism, empathy, motivation, overall quality. The questions are included in Supplementary Table 2.

## 3. Results

Quantitative analysis (Table 4) showed that the number of turns of dialogue was higher in coach-primed versus unprimed. Across both architectures, priming was associated with a significant boost in the rate of conversations ending in a positive user sentiment, the rate of question-asking by the coach LLM, and the use of coaching-related vocabulary.

**Table 4:** Quantitative assessment of unprimed versus coach-primed LLM conversations.

| Metric | Unprimed | Coach-primed | p value |
| --- | --- | --- | --- |
| Average length of LLM reply (# words ± S.D.) | 25.7 ± 6.5 | 23.7 ± 7.1 | 0.3 |
| Turns of conversation by user/LLM (# turns ± S.D.) | 3.1 ± 0.3 | 3.7 ± 0.7 | 0.2 |
| Conversations ending with positive user sentiment (%) | 30 | 60 | **0.02** |
| Conversations containing a question asked by LLM (%) | 0 | 30 | **0.03** |
| Conversations containing coaching-specific words used by LLM (%) | 40 | 80 | 0.08 |

To determine whether ratings for the primed and unprimed models differed from each other, we ran a series of linear mixed model analyses. These included a fixed effect for primed vs unprimed, and random effects for rater and prompt to account for non-independence of the observations. Regarding message content, the ratings of blinded reviewers were overall more favorable for the coach-primed LLMs. The (Table 5). Specifically, the coach-primed model was rated as significantly higher in terms of quality, providing actionable suggestions, and using realistic language. The ratings for the classifier re-ranked versus unprimed were less conclusive, but this may be because those ratings were based on a single statement response from the model rather than a full back-and-forth dialogue.

**Table 5:** Qualitative assessment of unprimed versus coach-primed LLM conversations based on the reviews of 8 adjudicators.

| Survey question (1, strong disagree 5, strong agree) | Unprimed | Coach-primed | p value |
| --- | --- | --- | --- |
| *Which conversation provides a better overall coaching experience (% , remainder unsure)* | 21 | 72 | - |
| *The quality of the coaching experience is high* | $3.42 \pm 0.88$ | $3.97 \pm 1.0$ | **<0.001** |
| *The coach provides concrete fitness strategies that are actionable to the user* | $3.61 \pm 1.1$ | $4.25 \pm 0.84$ | **<0.001** |
| *The coach provides motivation or encouragement to the user* | $3.68 \pm 1.1$ | $3.97 \pm 0.97$ | 0.095 |
| *The coach is empathetic toward the user's needs and challenges* | $3.51 \pm 1.0$ | $3.83 \pm 1.1$ | 0.123 |
| *The language used by the coach is realistic and appropriate for the setting* | $3.71 \pm 1.1$ | $4.10 \pm 1.0$ | **0.02** |

Tables 6 and 7 show the performance of the user and coach statement classifiers, including the size and label distribution in the train and test sets. The BERT-base model had 81% multi-class accuracy in accurately classifying the coach message as motivation, capability or opportunity.

**Table 6:**
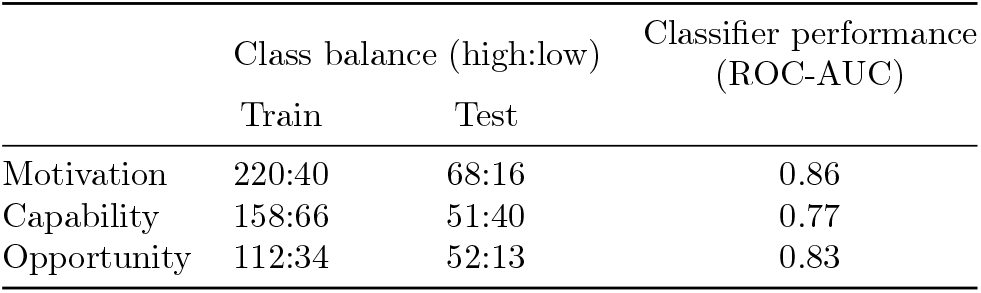
Class balance and model performance on C/O/M classification for user statements.

**Table 7:**
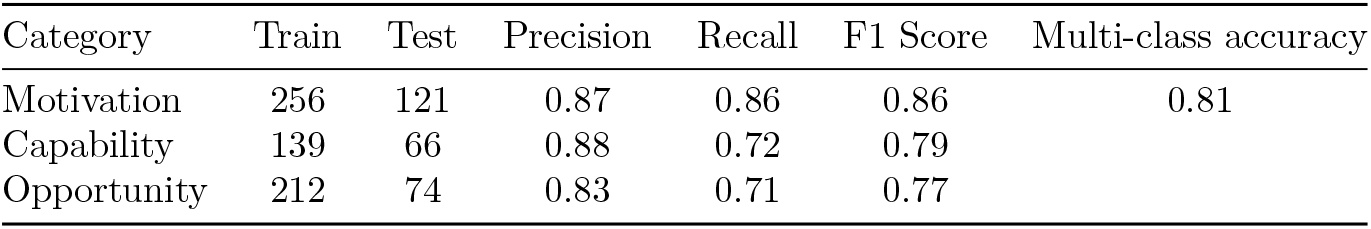
Model performance on C/O/M classification for coach statements.

To quantitatively evaluate the re-ranked response compared to the default response, 8 independent reviewers rated both the responses across several dimensions of activity coaching (Table 8). Based on Likert scale responses, the re-ranked answers were rated as more actionable [3.66*±*0.89 vs 2.88*±*0.85]; however the other attributes did not reach statistical significance.

**Table 8:** Qualitative review of coach-primed versus classifier re-ranked and oracle re-ranked dialogues.

| Survey question (1, strong disagree → 5, strong agree) | Unprimed | Coach-primed | Classifier Re-ranked | Oracle Re-ranked | p value<br>(Classifier re-ranked<br>vs Unprimed) |
| --- | --- | --- | --- | --- | --- |
| *The coach response provided concrete fitness strategies that are actionable* | 2.88 ± 0.85 | 3.22 ± 0.95 | 3.66 ± 0.89 | 4.02 ± 0.68 | **0.02** |
| *The coach response to user questions was in a realistic manner* | 3.02 ± 0.98 | 3.23 ± 0.97 | 3.59 ± 0.99 | 3.75 ± 0.80 | 0.18 |
| *The coach response provided motivation or encouragement to the user* | 3.05 ± 1.0 | 3.19 ± 0.85 | 3.75 ± 0.92 | 3.45 ± 1.05 | 0.36 |
| *The coach is empathetic toward the user's needs and challenges* | 2.94 ± 0.99 | 3.05 ± 0.94 | 3.56 ± 0.87 | 3.77 ± 0.83 | 0.47 |
| *The language used by the coach is realistic and appropriate for the setting* | 3.33 ± 0.87 | 3.48 ± 0.84 | 3.69 ± 0.87 | 3.78 ± 0.79 | 0.29 |
| **Average total score** | 3.04 ± 0.95 | 3.24 ± 1.36 | 3.65 ± 1.32 | 3.75 ± 0.84 | |

## 4. Discussion

This proof-of-concept study introduces two methods to infuse behavior science into LLM dialogue. We demonstrate that behavior science-based priming is a simple but effective strategy to tailor LLMs for activity coaching, with specific benefits in terms of actionability and the provision of concrete coaching advice. Additionally, post-hoc re-ranking of LLM responses based on behavior science principles can further enhance attributes such as perceived empathy.

BS priming yielded some significant boosts in various proxies for coaching quality. This trend was evident across both quantitative and qualitative metrics. Notably, coach phrase priming was associated with a higher number of conversational turns, a greater rate of question-asking, and more frequent use of coaching vocabulary. Manual review also judged coach phrase priming as providing significantly greater motivation and concrete coaching strategies versus the unprimed LLM. This suggests that BS priming may be an effective and accessible strategy for customising LLMs for various coaching scenarios.

A unique aspect of this work is the combination of priming with post-hoc re-ranking to enable knowledge infusion at multiple touchpoints. Interestingly, re-ranking resulted in significant incremental improvements in actionability, with upward trends in empathy, motivation and realism that did not meet statistical significance. We demonstrate this uplift both for a classifier-based re-ranking, which introduces error from mis-classification; and for oracle-based re-ranking, which showed a further marginal advantage over the former. Together, these results demonstrate the ability to stitch together multiple simple constraints as part of a hybrid knowledge infusion strategy. As LLMs become more pervasive in the coaching domain, this will be increasingly important.

Since Capability has marginally lower user statement classifier accuracy, it was wrongly identified as motivation in few cases of classifier based BeSci dialogue alignment LLM. This resulted in higher motivational character to classifier based LLM over Oracle LLM at the expense of lower empathy and actionability scores. This study has a number of limitations. First, the evaluation was predominantly based on simulated conversations with a single human interacting with the LLMs, which invariably introduces bias even in the presence of blinding. Future work could trial a similar evaluation with larger groups of users engaging in dialogue, as per [ref]. The rudimentary priming method used here could be extended, e.g. by more explicit instruction prompting or chain of thought prompting. The re-ranking method was limited in only focusing on a single user query and coach response. In reality, it is important to consistently align the coach responses to user need throughout a conversation and adapt as the dialogue unfolds. Methods such as reinforcement learning with human feedback can help to offer this adaptability (29). Finally, the behaviour model used was a simplistic one that conceptualizes user behaviour only along three axes - future studies could consider using more sophisticated behavior science frameworks, which may help to better target coach actions.

## 5. Conclusion

Knowledge infusion methods based on behavior science principles can be used to improve the quality of LLM-generated physical activity related conversations. The combination of coach phrase priming with re-ranking of LLM outputs offers optimal results in terms of manually-adjudicated actionability, empathy and overall coaching experience. These methods can help to constrain and guide LLMs in various coaching scenarios.

## Data Availability

Conversation queries are made available in supplementary code. The code is also made publicly available

https://github.com/fitllm/classifiers

