## Supplementary material for "Infusing behavior science into large language models for activity coaching"

### Section 1: Coach Phrase Priming sentence selection method

30 sentences crafted by 6 fitness experts were used for priming LLMs consisting of 10 sentences from capability, opportunity and motivation (COM) themes according to the COM-B model. These sentences were crafted to cater most common fitness coaching related queries experienced by the coaches. The handcrafted messages were provided as template conversations in the conversation tool for the WOO arm in the PACE study[ref]. The PACE algorithm selected a suitable template as response to user query according to the FBM model. The coaches were free to copy as is or modify the template or write their own response in the wizard of Oz setup.

The efficacy of the sentences used in priming were tested using the bleu score by matching the coach responses collected from the PACE study and output generated by the unprimed/primed LLMs. The Supplementary Table 1 describes the bleu match score LLM response with or without priming. The BLEU score (BiLingual Evaluation Understudy) is a metric for comparing similarity between a query and reference text based on matching n-grams. The sentences selected for the coach phrase priming have equal representation of motivation, opportunity and capability messages. These carefully selected sentences caused the LLMs output to have the highest match with the coach utterances in the PACE study as measured by the BLEU score.

| **Bleu Score** | **1-gram** | **2-gram** | **3-gram** | **4-gram** |
|---|---|---|---|---|
| Non Primed | 7.31 | 4.24 | 2.57 | 1.78 |
| Primed using randomly selected 30 coach sentences from PACE study | 17.89 | 8.52 | 5 | 3.34 |
| Primed with coach crafted 30 sentences for PACE study | 22.312 | 10.894 | 5.356 | 3.289 |

Supplementary Table 1 : BLEU match score to compare LLM primiring strategies to match human coach sentences.

The order of the 30 selected sentences used in priming influences the LLMs output as shown in the previous works[ref]. When the sentences used to prime are batched by the theme (COM), then the output is biased towards the last theme. We randomly order the sentences mixing all three themes but the order is kept consistent when priming for each conversation for reproducibility. Priming based on one of the COM-B themes narrows the LLM output and the quality as measured qualitatively. Since the user query can span across a wide range of fitness issues and straddle multiple COM themes, we do not prime the LLMs with any single predetermined themed sentences. Rather, a custom re-ranking algorithm described later is used for tighter control of the COM-theme primed LLMs output to cater to the specific user query.

### Section 2 : LLM based conversation evaluation tool and methodology

A conversational simulator (Supplementary figure 1) to display the coach and user conversation session was developed  to test the efficacy of the proposed LLM enhancements. Each conversation session in the simulator consists of a series of dialogue exchanges between user and LLM to resolve a specific user generated fitness issue. The conversation tool prompts the following message to initiate the conversation “Hey John, It's time for your morning walk.” A preselected user query is given to LLM which sets the conversation topic on a fitness related issue. 8 independent raters assess the quality of the LLMs response conditioned on the user query.  The 9 user queries that were selected to test the LLM response as shown in supplementary table 2. These queries were not used in the training of user query classifiers. Three user queries from each of the COM themes were identified from various points in the course of the 21 day PACE study. These queries were the most commonly occurring user initiated conversations for each of the themes from the PACE study. The participants in the pace study were free to pick any walking related questions to ask the coaches. This evaluation strategy allows testing the LLM response on important user queries which may arise after multiple days of interaction with coaching assistants which is not possible if LLM are evaluated by having a one off conversation with a new participant each time. Also, having independent annotators to rate the conversation session helps reduce bias and also scale the number of evaluations to statistically significant numbers. Any follow up questions to the LLM response were added appropriately to continue the conversation on the original topic until a logical end is reached.

| A | |
|---|---|
| Coach | Hey John, It's time for your morning walk. |
| Human | Hi Coach - Yes, it is. I would more inclined to go if it could be a bit more fun. |
| Coach | That's a great attitude, John! Let's see if we can make your walk more fun. Maybe you could walk through a park or by a river. Second, let's try listening to some music or an audio book while you walk. Finally, let's see if we can get a friend or family member to join you on your walk. |
| Human | That sounds like a great plan! I'm going to go ask my wife if she wants to join me on my walk today. Thanks for the motivation, Coach! |

| B | |
|---|---|
| Coach | Hey John, It's time for your morning walk. |
| Human | Hi Coach - Yes, it is. I would more inclined to go if it could be a bit more fun. |
| Coach | Well, let's make it more fun then! Why don't we listen to some music or a podcast that you enjoy while we walk? |
| Human | That could probably help. But I still get bored alone. |
| Coach | How about we walk with a friend then? |
| Human | Maybe that could make walking less boring and more fun. |
| Coach | Let's try it and see! |
| Human | Thank you for the suggestion coach. |
| Coach | You're welcome! Let me know how it goes. |

| ID | Question | A | B |
|---|---|---|---|
| 1 | Which conversation provides a better overall coaching experience - A, B or unsure | B | |
| 3 | The coach provides concrete fitness strategies that are actionable to the user (1 - strongly disagree ⟶ 5- strongly agree) | 5 | 5 |
| 4 | The coach responds to user questions in a realistic manner (1 - strongly disagree ⟶ 5- strongly agree) | 2 | 4 |
| 5 | The coach provides motivation or encouragement to the user (1 - strongly disagree ⟶ 5- strongly agree) | 4 | 4 |
| 6 | The coach is empathetic toward the user's needs and challenges (1 - strongly disagree ⟶ 5- strongly agree) | 4 | 4 |
| 7 | The language used by the coach is realistic and appropriate for the setting (1 - strongly disagree ⟶ 5- strongly agree) | 2 | 4 |

Supplementary figure 1 : LLM Conversation rating tool used by annotators

8 raters assessed the LLM response  qualitatively for a given user query by answering 8 survey questions.  The raters had an equal mix of age, gender and demographics without any prior fitness coaching experience. Raters were not incentivised for this exercise and were blinded to the LLM variation which generated the automated response and to the LLM

technology. Raters used survey forms to submit the rating for each set of user - LLM conversation and were free to change the ratings at any time during the exercise.

| Index | Category | User query | Days |
|---|---|---|---|
| 1 | **Motivation** | I was able to meet my goal of step count in yesterday's walk, but sometimes it is hard to meet the goal. | 7 |
| 2 | **Motivation** | I would be more inclined to go walking if it could be a bit more fun. | 5,8 |
| 3 | **Motivation** | I am busy today and not sure of all the benefits of daily walking on health and well-being. | 2,4 |
| 4 | **Capability** | Hi coach, unfortunately I have injured myself. I can't walk today. | 5,11 |
| 5 | **Capability** | I really don't think it is the best use of time. I just don't think it would be worth it. | 9 |
| 6 | **Capability** | Hi coach, it is raining outside. I do not want to walk in the rain and I do not have a treadmill. Is there anything else I can do? | 6,2 |
| 7 | **Opportunity** | I am super busy with work today.I have chores to do in the morning and work meetings after that. | 1,7,5 |
| 8 | **Opportunity** | Hi coach, sure but I am not able to get it in my routine. How can I plan better ? | 4,2,8 |
| 9 | **Opportunity** | Hi Coach, 30 minutes would be a lot. Is there any other alternative you could suggest? | 8,8 |

Supplementary Table 2 : User queries across COM themes selected for LLM evaluation. Days column represent the days when the query was asked by different users to coaches in the PACE study

### Section 3. Coach response & User query classifier

To provide context relevant responses with right behavior science backed construct, it is important to recognize the topic queried by the user on walking and respond back on the same theme to address them and resolve the issue to enable walk more. Though LLMs are trained on a wide corpus and do reasonably well, there is potential to improve further, since LLMs do not explicitly understand behavior science constructs like motivation, capability, & opportunity nor have any in-built coaching specific classifier to determine user query topic. Given a user query sentence, the *user-query* classifier assigns high vs low value for each of the motivation, capability & opportunity (COM) dimensions. The COM-B model is used to pick the appropriate theme for coach response based on output values for COM dimensions as shown in figure 2 in methods. COM-B policy prioritizes addressing the category with low value and using the priority order : capability, opportunity and motivation to resolve any conflicts. PACE study showed ~95% of the user query could be classified as either high/low on atleast one of COM with confidence. Post action selection, the Reranking algorithm is used to modify LLM response to suit the selected COM theme.

| Capability | Opportunity | Motivation | COM-B Action |
|---|---|---|---|
| Low | High/Low | High/Low | Boost Capability |
| High | Low | High/Low | Boost Opportunity |
| High | High | High/Low | Boost Motivation |

Supplementary Table 3 : COM-B policy to select nudge theme based on COM values derived from user query classifier

The Reranking method builds on top of primed LLM which generates richer conversation responses. LLMs generate many candidates and stack rank using in-built logic developed in the training process. They fire the top-1 as the response to the user query. LLMs can be personalized further for fitness for coaching by recognizing the user query topic/theme and using it to re-rank the generated sentences by primed LLMs to match the user query theme based on COM-B policy. For example, if the user seeks alternatives to walking during rain, ReRanked LLMs recognize the user theme as lack of capability, and use the information to re-rank the capability enhancing messages relative to motivation and opportunity as suggested by the COM-B algorithm. Success of the method hinges on developing sufficiently accurate NLP based user query and LLM response classifiers to identify one of the COM themes for each input sentence.

### Section 4 : Summary of PACE study and adaptation to FIT-LLM work

The PACE coaching conversation tool was built to test the efficacy of automated conversational assistants in comparison to fitness experts to assist participants to improve walking habits through text based conversations on digital platforms as described in the paper[ref]. The Wizard of Oz (WOO) or treatment group had coaching conversations selected using COM-B policy to match user query. The PACE tool consisted of the coaching message repository crafted by the fitness experts and the COM-B algorithm to select the theme of the response to the user query. Coaches are free to use the suggested message by the tool, edit them or craft a new response on their own. Coaches went with the PACE tool suggested response in 80%+ cases and the efficacy of the WOO arm was comparable to fitness coach led control arm in both the qualitative and quantitative metrics discussed in the PACE paper. 520 conversations were generated using the PACE tool to assist 16 participants for the duration of 14 days on a wide variety of fitness related topics. A total of 6 independent annotators labeled these conversations as one of Motivation, Capability and Opportunity.A conversation session is usually on a single fitness topic but may have many pairs of user &

coach dialogue exchanges leading to multiple training examples. This serves as a train & test set for respective classifiers. These conversations were used to train and evaluate coach response and user query theme classifier. Further, user queries with pre-defined criteria were selected to evaluate the LLMs post priming and re-ranking. This method led to selection of the most commonly asked user queries to test efficacy of LLMs on a wide variety of fitness related topics. The BERT [ref] is used as a base model to fine tune for user query and coach response COM classifiers. Weights are not shared between the models and are trained independently. The last classification layer is re-trained with cross entropy loss to develop a 3-way classifier to classify any input sentence as one of C/O/M.

### Section 5 : Behavior Science Infusion to LLM framework pipeline for user query

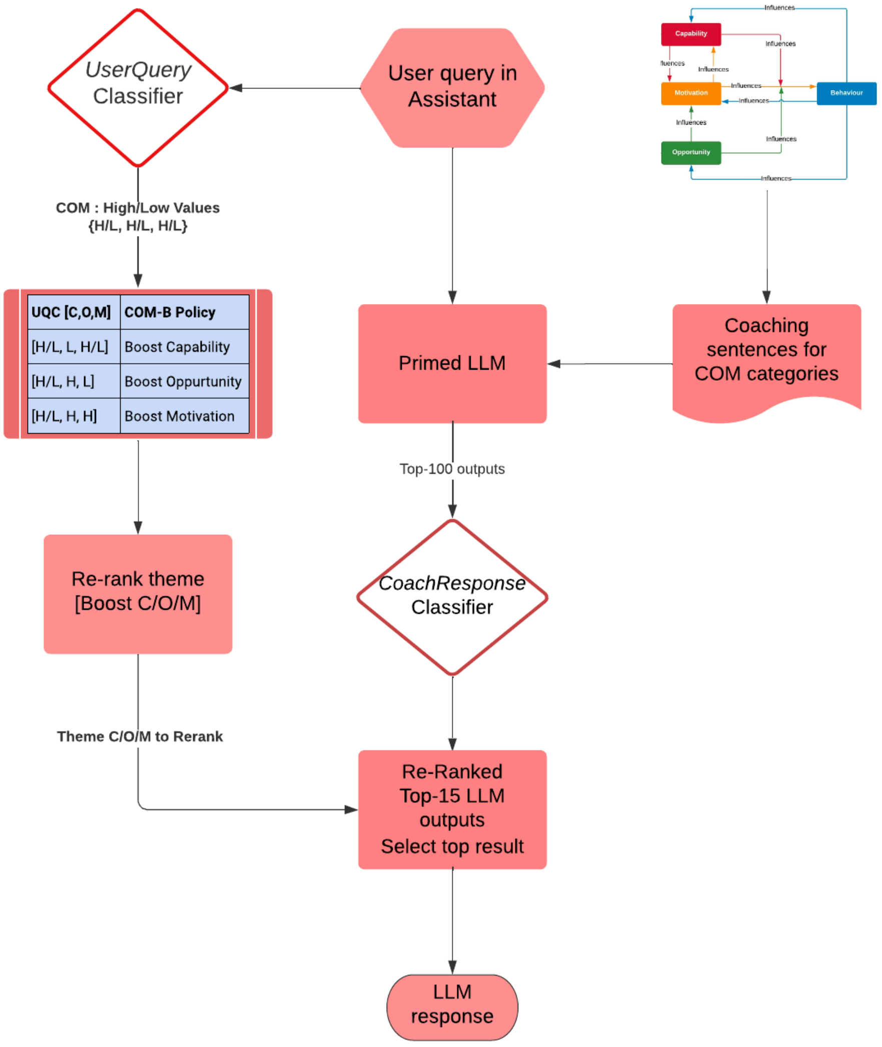


Supplementary Figure 2: Priming and BeSci infusion pipeline in Fit-LLM for user query input
